# Broncho-alveolar inflammation in COVID-19 patients: a correlation with clinical outcome

**DOI:** 10.1101/2020.07.17.20155978

**Authors:** Laura Pandolfi, Tommaso Fossali, Vanessa Frangipane, Sara Bozzini, Monica Morosini, Maura D’Amato, Sara Lettieri, Mario Urtis, Alessandro Di Toro, Laura Saracino, Elena Percivalle, Stefano Tomaselli, Lorenzo Cavagna, Emanuela Cova, Francesco Mojoli, Paola Bergomi, Davide Ottolina, Daniele Lilleri, Angelo Guido Corsico, Eloisa Arbustini, Riccardo Colombo, Federica Meloni

## Abstract

Severe acute respiratory syndrome coronavirus 2 (SARS-CoV-2) rapidly reached pandemic proportions. We conducted a prospective study to assess deep lung inflammatory status in patients with moderate to severe COVID-19.

Diagnostic bronchoalveolar lavage (BAL) was performed in 33 adult patients with SARS-CoV-2 infection by real-time PCR on nasopharyngeal swab admitted to the Intensive care unit (ICU) (n=28) and to the Intermediate Medicine Ward (IMW) (n=5). We analyze the differential cell count, ultrastructure of cells and Interleukin(IL)6, 8 and 10 levels.

ICU patients showed a marked increase in neutrophils (72%, 60-81), lower lymphocyte (8%, 4-12) and macrophages fractions (17%, 11-27) compared to IMW patients (3%, 2-17, 15%, 6-26 and 74%, 58-90, respectively) (p<0.01). Ultrastructural study from ICU patients showed viral-like particles in cytopathic mononuclear cells however extensive cytopathic damage in all cell lineages. Immunostaining with anti-viral capsid and spike antibodies specifically immunoreacted with BAL cells, mostly cytopathic ones. IL6 and IL8 were significantly higher in ICU patients than in IMW (IL6 p<0.01, IL8 p<0.0001), and also in patients who did not survive (IL6 p < 0.05, IL8 p = 0.05 *vs*. survivors). IL10 did not show a significant variation between groups. Dividing patients by treatment received, lower BAL concentrations of IL6 were found in patients treated with steroids as compared to those treated with tocilizumab (p<0.1) or antivirals (p<0.05).

Alveolitis, associated with COVID-19, is mainly sustained by innate effectors which showed features of extensive activation. The burden of pro-inflammatory cytokines IL6 and IL8 in the broncho-alveolar environment is associated with clinical outcome.

## Introduction

Severe acute respiratory syndrome coronavirus 2 (SARS-CoV-2), detected in Wuhan (China) in December 2019 [1], spread rapidly around the world, reaching pandemic proportions in March 2020. In Italy, from February up to June 17^th^, 237,500 SARS-CoV-2-positive adults have been documented with 34,405 deceased patients [2]. Since December 2019, researchers and clinicians have been trying to find out the pathogenic features of the new disease caused by SARS-CoV-2, named COVID-19, in order to develop an effective therapy. The typical clinical manifestation is severe interstitial pneumonia with hyperactivation of the inflammatory cascade and progression to acute respiratory distress syndrome (ARDS) in some cases [3-5]. As of yet, most research papers have focused on the inflammatory status at the plasma level of COVID-19 patients [4-8]. However, because the main target organ is the lung, it is crucial to understand the inflammatory status at the deep lung level during different stages of the infection. Currently, limited data are available about alveolar inflammatory status in COVID-19 patients because of concerns in relation to using bronchoscopy to avoid aerosol generation. A transcriptomic analysis was conducted by Xiong et al. on bronchoalveolar lavage (BAL) samples obtained from a small number of COVID-19 patients and measured the expression levels of cytokines and chemokines [9]. Another report comparing three patients with moderate COVID-19 and six patients with severe infection through a single-cell RNA sequencing (scRNA-seq) on BAL cells revealed aberrant macrophage activation and the formation of tissue-resident CD8+ T cell in the lung of mild symptom patients [10].

Herein, we aim to investigate the deep lung inflammatory status via the analysis of BAL cells and the cytokine profile in patients with severe COVID-19 (N = 28), comparing them with patients admitted to the intermediate medicine ward (IMW) (N = 5). Patients were enrolled at the Luigi Sacco Hospital (Milan, Italy) and the IRCCS Policlinico San Matteo Foundation Hospital (Pavia, Italy), two COVID-19 hospitals situated at the epicenter of the Italian epidemic outbreak.

## Methods

### Patients

This study included 33 adults positive for SARS-CoV-2 infection, diagnosed by real-time PCR on nasopharyngeal swab. 28 patients with severe ARDS requiring intensive respiratory support (mechanical ventilation or extracorporeal support) were admitted to the Intensive Care Unit (ICU) at the Luigi Sacco Hospital (Milan, Italy) (N = 25), or the IRCCS Policlinico San Matteo Foundation Hospital (Pavia, Italy) (N=3) (henceforth referred as ICU pts). The remaining 5 patients had moderate pneumonia with PaO_2_/FiO_2_ ratio > 250 and were admitted to the Intermediate Medicine ward (IMW) (N = 5) of the IRCCS Policlinico San Matteo Foundation.

Bronchoscopies were performed at both centers for diagnostic purposes in order to investigate the presence of other co-infections.

Research and data collection protocols were approved by the Institutional Review Boards (Comitato Etico di Area 1) (prot. 20100005334) and by IRCCS Policlinico San Matteo Foundation Hospital (prot. 20200046007), written informed consent was provided by patients when possible (IMW and conscious ICU patients) and was waived in all other cases.

### Clinical variables included in the analysis

Demographic and clinical characteristics of the enrolled patients were recorded in a dedicated database. Failure of a trial with continuous positive airway pressure via a helmet was the indication for admission in ICU. Failure was defined as respiratory rate >30 breaths per minute and PaO_2_ to FiO_2_ ratio <150, or respiratory acidosis with pH<7.36 and PaCO_2_ >50 mmHg, or agitation, or confusion. Laboratory findings, including complete blood count, basic metabolic panel, indexes of inflammation and hypercoagulability, levels of serum Interleukin(IL)6 (when available), and blood gas analysis were recorded for the duration of hospitalization. Ventilator settings and therapies such as corticosteroids, antiviral drugs, IL-6 antagonists or hydroxychloroquine or other antivirals were also recorded.

### BAL collection

BAL was collected from mechanically ventilated ICU patients according to the clinical requirement through disposable bronchoscope aScope™ 4 (Ambu A/S, Baltorpbakken, Denmark). Six patients underwent repeated bronchoscopies. Therefore, the number of overall BAL samples were 45. When disposable bronchoscopes were not available, we followed the standard High-Level Disinfection for the utilization of re-usable bronchoscopes. Security procedures were strictly observed, and only essential personnel were involved in endoscopic exams. The Personal Protective Equipment (PPE) included water-resistant gowns, gloves, respiratory protection (FFP3 mask), and eye protection. BAL specimens were managed in a biosafety level 3 laboratory until inactivation. BALs were centrifuged at 400 g for 10 min at room temperature, inactivated with a 0.2% SDS and 0.1% Tween-20 solution followed by 65 °C for 15 minutes. BAL supernatants were then stored at -20°C until analysis. Cell pellets were fixed, then stained with Papanicolaou to analyze differential cell count or used for ultrastructural analysis.

### Ultrastructural analysis and immunostaining

BAL samples were fixed with Karnovsky’s fixative, treated with 1.5% OsO_4_ in 0.2 mol l^-1^ cacodylate buffer (pH 7.3), dehydrated and embedded in Epon-Araldite resin. Ultrathin sections were stained with lead citrate and uranyl acetate and examined with a electron microscope (JEOL JEM-1011). SARS-CoV-2 infected VERO E6 cells at 48 and 72 hours were used as positive controls.

The immunostaining of paraffin-embedded sections was done using SARS-CoV-2 (2019-nCoV) Nucleoprotein / NP Antibody, Rabbit MAb (Sino Biological, Catalog number: 40143-R019) – Dilution 1:1000 and SARS-CoV-2 (2019-nCoV) Spike Antibody, Rabbit MAb (Sino Biological, Catalog number: 40150-R007) - Dilution 1:400.

### ELISA Assays

To quantify IL8, IL10 and IL6, we used the SimpleStep ELISA® kit (Abcam, Cambridge, UK). Briefly, 50 μl of each sample was added to ELISA kit wells with the addition of 50 μl antibody cocktail. After 1 hour at room temperature on a plate shaker, plates were washed three times to eliminate the unbound antibody. Substrate (100 μl) was incubated for 10 min in the dark at room temperature on a plate shaker, followed by 100 μl stop solution to read the absorbance at 450 nm. For IL8 and IL6, samples were diluted (from 1:10 to 1:1000). To assess that the inactivation protocol that we adopted did not alter the quantification of citokines, we firstly tested non COVID-19 infected BAL fluids with or without inactivation solution, obtaining the same quantification of all three citokines (data not shown).

### Statistical analysis

Comparison of cell counts between ICU and IMW was analyzed by multiple t-test followed by the Sidak method. Analysis of cytokines between ICU vs. IMW or survivors and non-survivors were carried out using the Mann-Whitney test. With regard to the analysis of cytokines according to treatment strategies we used the Kruskal-Wallis test followed by Dunn’s multiple comparison test. Association between variables was assessed with Spearman’s correlation. Statistical significance was defined as p ≤ 0.05. All data are represented as median (interquartile range – IQR). Data were statistically analyzed with Graphpad Prism version 8.4.1.

## Results

From March 7 to April 30, a total of 45 BAL samples were collected from 5 IMW and 28 ICU patients with proven COVID-19 infection. Patients characteristics are shown in Table 1, and data concerning administered treatment in Table 2. All patients from the IMW ward survived. Significant differences regarding laboratory abnormalities among ICU and IMW patients, when present, are shown in Table 1.

**Table 1.**
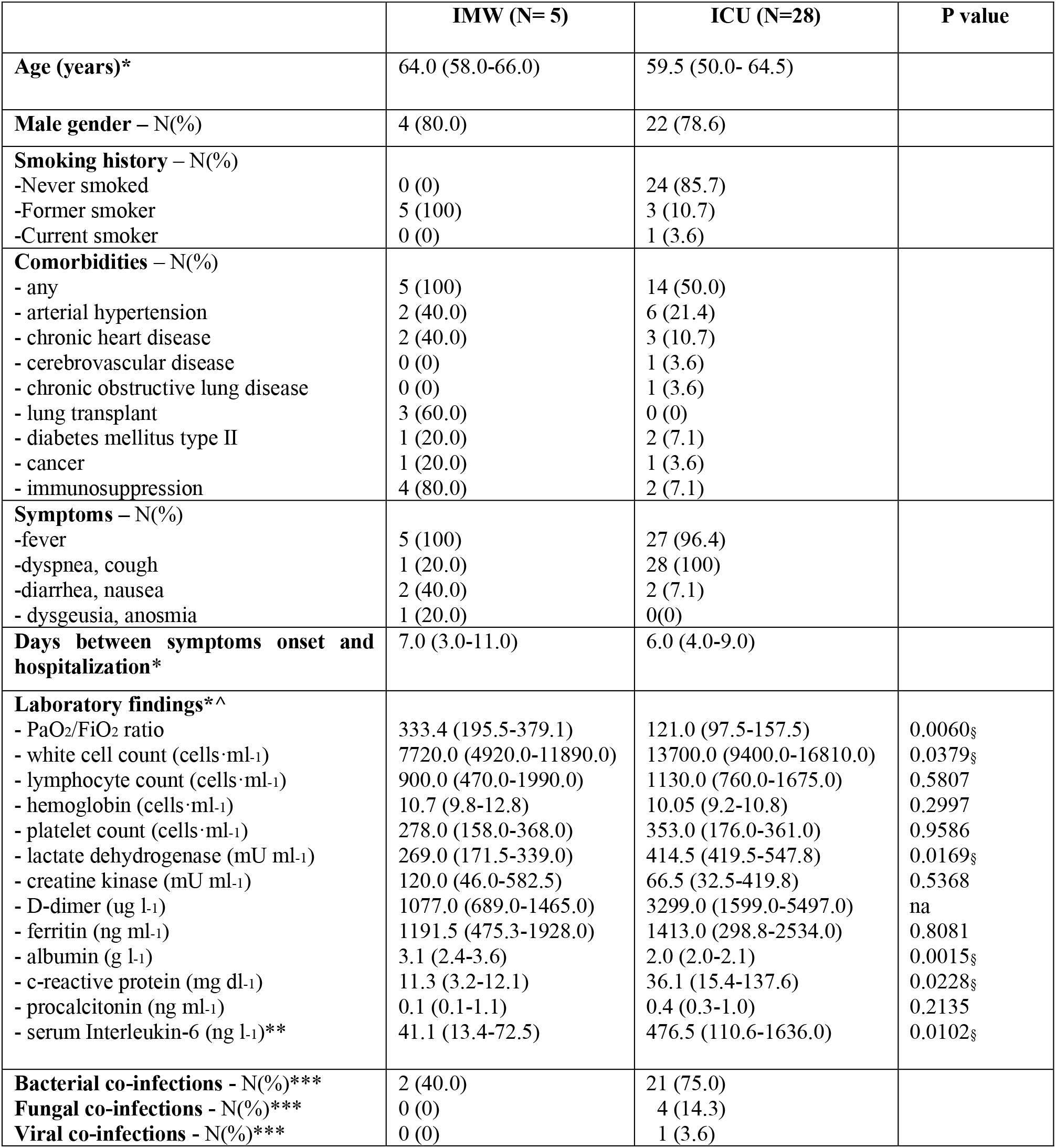
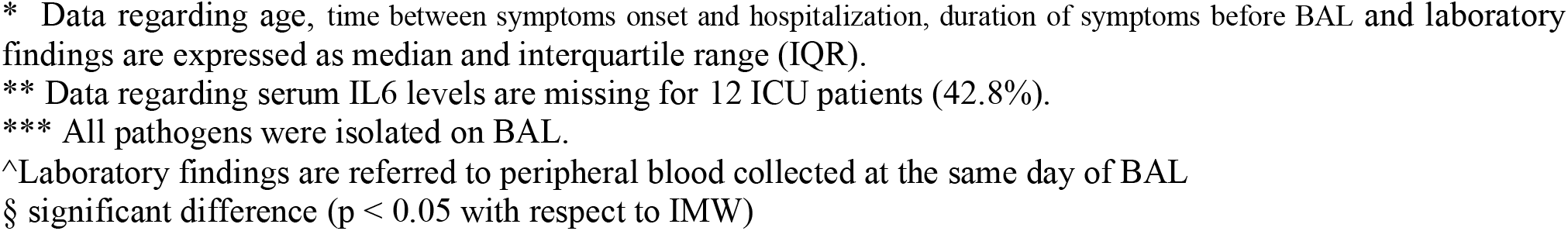
Demographic characteristics of the studied population and laboratory findings at admission

|  | IMW (N= 5) | ICU (N=28) | P value |
| --- | --- | --- | --- |
| **Age (years)\*** | 64.0 (58.0-66.0) | 59.5 (50.0- 64.5) |  |
| **Male gender – N(%)** | 4 (80.0) | 22 (78.6) |  |
| **Smoking history – N(%)** |  |  |  |
| -Never smoked | 0 (0) | 24 (85.7) |  |
| -Former smoker | 5 (100) | 3 (10.7) |  |
| -Current smoker | 0 (0) | 1 (3.6) |  |
| **Comorbidities – N(%)** |  |  |  |
| - any | 5 (100) | 14 (50.0) |  |
| - arterial hypertension | 2 (40.0) | 6 (21.4) |  |
| - chronic heart disease | 2 (40.0) | 3 (10.7) |  |
| - cerebrovascular disease | 0 (0) | 1 (3.6) |  |
| - chronic obstructive lung disease | 0 (0) | 1 (3.6) |  |
| - lung transplant | 3 (60.0) | 0 (0) |  |
| - diabetes mellitus type II | 1 (20.0) | 2 (7.1) |  |
| - cancer | 1 (20.0) | 1 (3.6) |  |
| - immunosuppression | 4 (80.0) | 2 (7.1) |  |
| **Symptoms – N(%)** |  |  |  |
| -fever | 5 (100) | 27 (96.4) |  |
| -dyspnea, cough | 1 (20.0) | 28 (100) |  |
| -diarrhea, nausea | 2 (40.0) | 2 (7.1) |  |
| - dysgeusia, anosmia | 1 (20.0) | 0(0) |  |
| **Days between symptoms onset and hospitalization\*** | 7.0 (3.0-11.0) | 6.0 (4.0-9.0) |  |
| **Laboratory findings\*^** |  |  |  |
| - PaO<sub>2</sub>/FiO<sub>2</sub> ratio | 333.4 (195.5-379.1) | 121.0 (97.5-157.5) | 0.0060§ |
| - white cell count (cells·ml<sup>-1</sup>) | 7720.0 (4920.0-11890.0) | 13700.0 (9400.0-16810.0) | 0.0379§ |
| - lymphocyte count (cells·ml<sup>-1</sup>) | 900.0 (470.0-1990.0) | 1130.0 (760.0-1675.0) | 0.5807 |
| - hemoglobin (cells·ml<sup>-1</sup>) | 10.7 (9.8-12.8) | 10.05 (9.2-10.8) | 0.2997 |
| - platelet count (cells·ml<sup>-1</sup>) | 278.0 (158.0-368.0) | 353.0 (176.0-361.0) | 0.9586 |
| - lactate dehydrogenase (mU ml<sup>-1</sup>) | 269.0 (171.5-339.0) | 414.5 (419.5-547.8) | 0.0169§ |
| - creatine kinase (mU ml<sup>-1</sup>) | 120.0 (46.0-582.5) | 66.5 (32.5-419.8) | 0.5368 |
| - D-dimer (ug l<sup>-1</sup>) | 1077.0 (689.0-1465.0) | 3299.0 (1599.0-5497.0) | na |
| - ferritin (ng ml<sup>-1</sup>) | 1191.5 (475.3-1928.0) | 1413.0 (298.8-2534.0) | 0.8081 |
| - albumin (g l<sup>-1</sup>) | 3.1 (2.4-3.6) | 2.0 (2.0-2.1) | 0.0015§ |
| - c-reactive protein (mg dl<sup>-1</sup>) | 11.3 (3.2-12.1) | 36.1 (15.4-137.6) | 0.0228§ |
| - procalcitonin (ng ml<sup>-1</sup>) | 0.1 (0.1-1.1) | 0.4 (0.3-1.0) | 0.2135 |
| - serum Interleukin-6 (ng l<sup>-1</sup>)** | 41.1 (13.4-72.5) | 476.5 (110.6-1636.0) | 0.0102§ |
| **Bacterial co-infections - N(%)\*\*\*** | 2 (40.0) | 21 (75.0) |  |
| **Fungal co-infections - N(%)\*\*\*** | 0 (0) | 4 (14.3) |  |
| **Viral co-infections - N(%)\*\*\*** | 0 (0) | 1 (3.6) |  |

\* Data regarding age, time between symptoms onset and hospitalization, duration of symptoms before BAL and laboratory findings are expressed as median and interquartile range (IQR).
\*\* Data regarding serum IL6 levels are missing for 12 ICU patients (42.8%).
\*\*\* All pathogens were isolated on BAL.
^Laboratory findings are referred to peripheral blood collected at the same day of BAL
§ significant difference ( $p < 0.05$ with respect to IMW)

**Table 2.** Treatment characteristics in ICU patients.

| TREATMENT | SURVIVORS<br>(N=16) | NON-SURVIVORS<br>(N=12) |
| --- | --- | --- |
| **Tocilizumab N(%)** | 4 (25) | 4 (25) |
| **Remdesivir N(%)** | 3 (18.8) | 4 (5) |
| **Hydroxychloroquine N(%)** | 12 (75) | 12 (100) |
| **Ritonavir/Lopinavir N(%)\*** | 7 (50) | 9 (75) |
| **Corticosteroids (0.3-0.5 mg·Kg<sup>-1</sup> body weight) N(%)\*\*** |  |  |
| - dexamethasone | 4 (25) | 5(42) |
| - methylprednisolone | 0 (0) | 1(8.3) |
| - hydrocortisone | 1 (6.25) | 1(8.3) |
| **Mechanical ventilation\*\*\*** |  |  |
| -Vt/IBW — ml Kg<sup>-1</sup>**** | 7.85(7.6-8.85) | 6.4 (6.1-8) |
| -PEEP — cmH<sub>2</sub>O | 11(8-13.25) | 12 (8-12) |

\* Data regarding the administration of Ritonavir/Lopinavir are missing for two patients who died and two who survived (14.8%)
\*\* Data regarding the administration of steroids is missing for one dead patient (3.70%)
\*\*\* Data regarding ventilation parameters are missing of two dead patients (7.41%)
\*\*\*\*Vt/IBW= Tidal volume/ideal body weight calculated using the gender-specific Acute Respiratory Distress Syndrome Network (ARDSnet) formulas [11].

Firstly, we assessed BAL cell differentials in ICU patients who presented a median of total cells of 0.25 × 10^5^ ml^-1^ (0.15 – 0.52) and in IMW patients with a median of total cells 0.3 × 10^5^ ml^-1^ (0.15 – 0.52). A different leukocyte profile between ICU (Fig. S1a) and IMW (Fig. S1b) patients was observed. In particular, ICU patients showed a marked prevalence of neutrophils (72%, 60-81) with a lesser extent of macrophages (17%, 11-27) and lymphocytes (8%, 4-12) as compared to IMW patients, who had median macrophages at 74% (58-90) lymphocytes at 15% (6-26) and neutrophils at 3% (2-17) (p<0.01).

IMW patients presented a significant decrease of macrophages (p<0.05) and a slight, but not significant (p=0.26), increase of lymphocyte and neutrophils with respect to normal reference values from our lab [12]. Dividing all patient samples into survivors and non-survivors, a significant difference was observed only for lymphocytes (Fig. S1e, p<0.01), but not for macrophages and neutrophils (Fig. S1c and S1d), although there was a trend towards a lower macrophage fraction among non-survivors.

Ultrastructural study of ICU BAL samples showed many cytopathic cells, with loss of integrity of cell membranes and cytoplasm vacuolization (Fig.1a-d). Mononuclear cells and neutrophils were detected with morphological features of activation (Fig. 2a and b). Viral particles, both single and in small clusters, were identified in numerous cells, particularly in cytopathic epithelial cells and mononuclear cells, with morphology (spikes) and size (80-120 nm) consisting with coronavirus (Fig. 1a-d and S2), confirmed by immunohistochemistry (Fig. 1e,f and 2d). Infected VERO E6 cells used as positive control showed large amounts of clustered and single viral particles clustered (Fig. S3).

**Figure 1.**
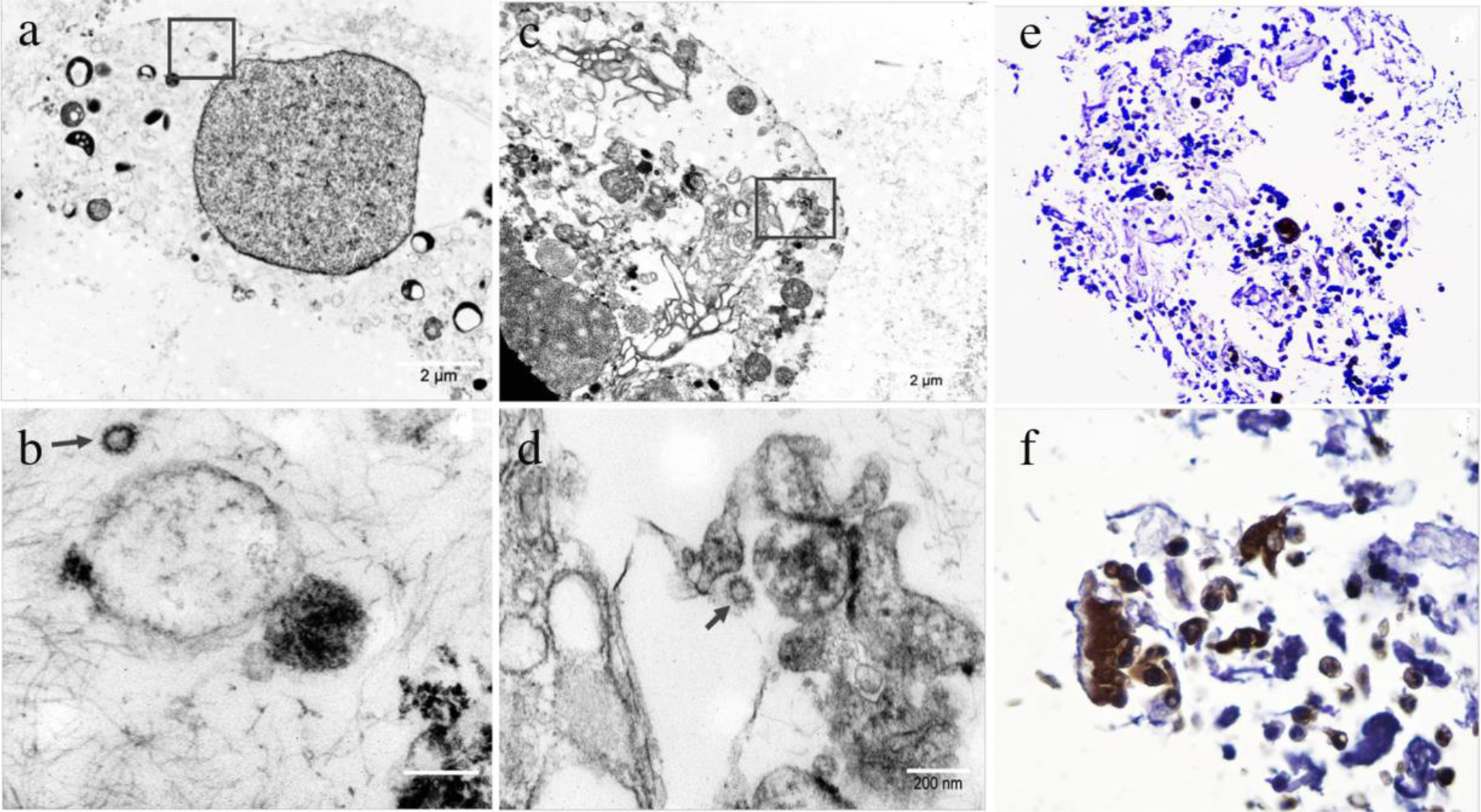
Severely cytopathic cells in BAL sample of ICU patient. (a) Squared area is enlarged in (b) that shows isolated viral particles in the cytoplasms; (c) shows a similar severe cytopathic cell: the squared area is enlarged in (d) with isolated viral particles. (e) BAL cells immunoreacting with anti-spike antibodies were variably represented in BAL samples, including (f) ciliated epithelia. Scale bar = 2 µm and 200 nm.

**Figure 2.**
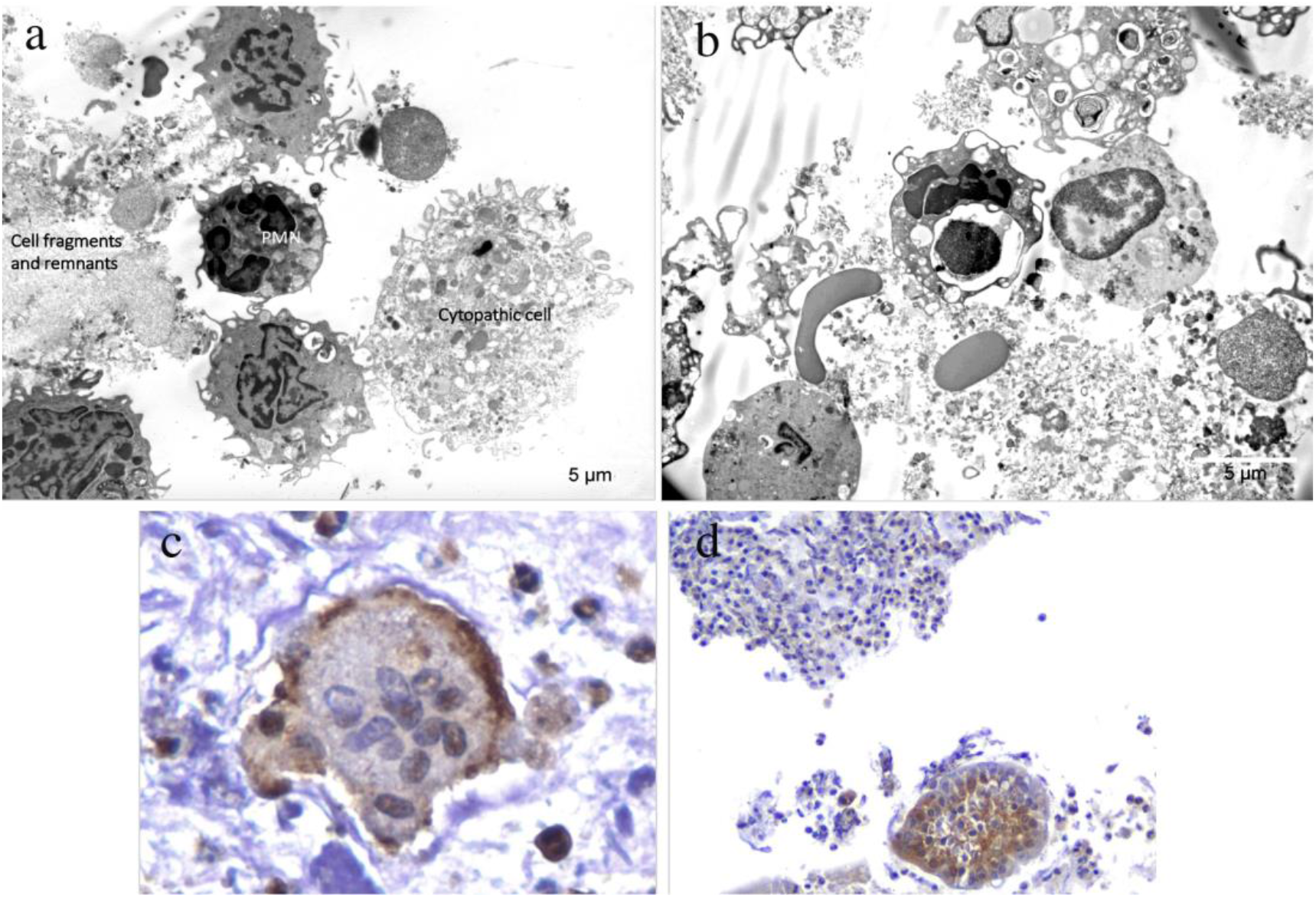
(a-b) Electron micrographs showing both (a) cytopathic cells and still morphologically preserved mononuclear cells and (b) extensive cytopathy involving all cells. (c) the SARS-CoV-2–anti-spike antibodies immunoreactive multinucleated cells in the BAL sample of one of the ICU patients; (d) flaps of epithelial cells and inflammatory cells immunoreacting with anti-spike antibodies. PMN = polymorphonuclear leukocyte. Scale bar = 5 µm

The following cytokines: IL6, IL8 and IL10 were quantified on cell-free BAL by ELISA. Figure 3a and 3b show the different distribution of three BAL cytokine levels between ICU (Fig. 3a) and IMW patients (Fig. 3b). Pro-inflammatory cytokines, IL6 and IL8, were significantly higher in ICU than in IMW patients (IL6 p<0.01 and IL8 p<0.0001), while IL10 did not show a significant difference between groups. When analyzing cytokines according to outcome, higher IL6 (Fig. 3c, p<0.05) and IL8 (Fig. 3d, p=0.05) BAL levels were detectable in non-survivors with respect to survivors, with no difference for IL10 (Fig. 3e).

**Figure 3.**
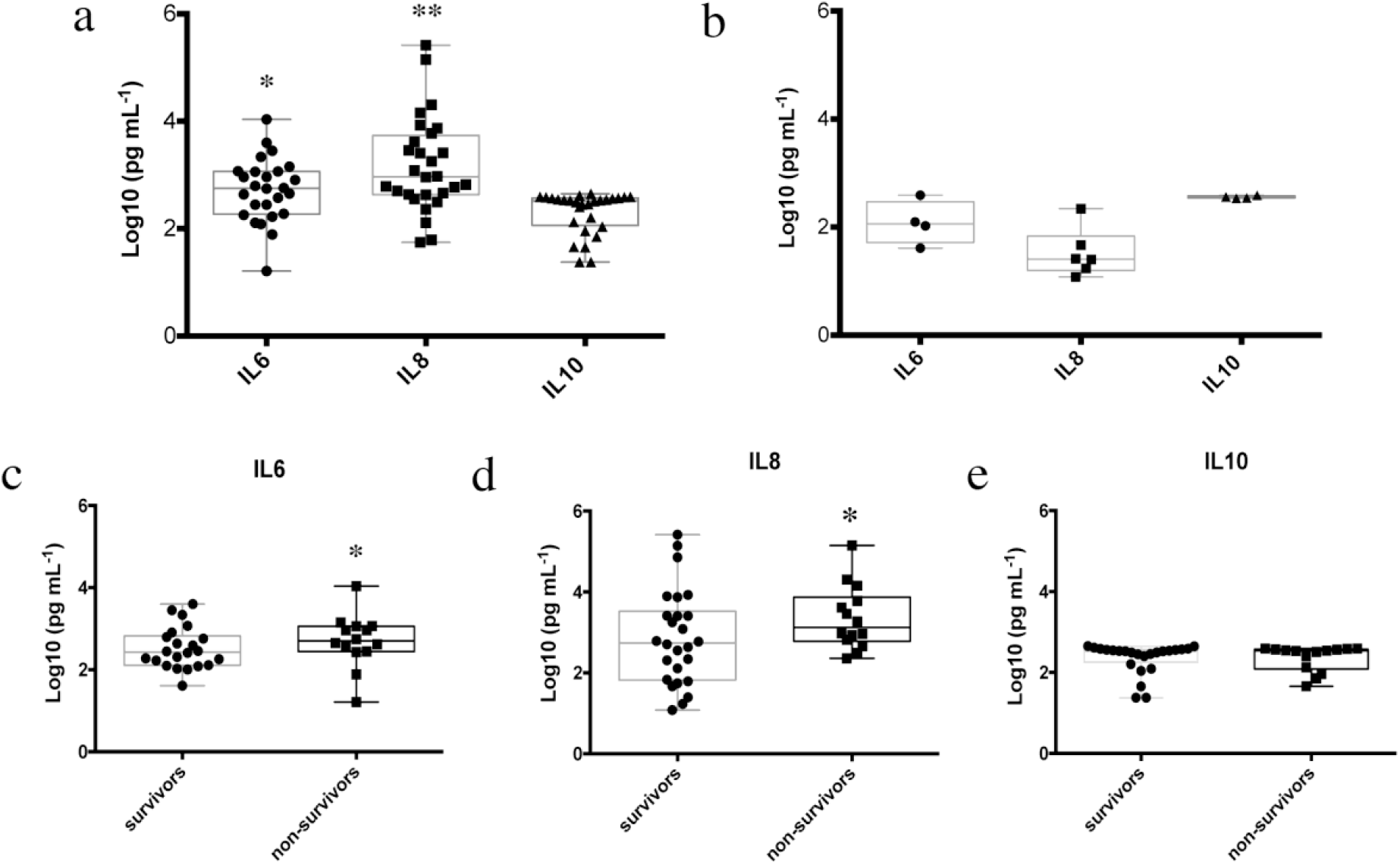
Distributions of cytokine values quantified in BAL of (a) ICU patients and (b) IMW patients (repeated BAL samples in the same patients have been excluded). *p<0.01 vs. IMW; **p<0.001 vs. IMW. (c-e) Quantified cytokines divided in survivors and non-survivors. *p ≤ 0.05 vs. survivors. Data has been transformed as Log10(pg ml^-1^) and represented as median (IQR).

Variations of cytokine BAL levels according to treatment strategies were also evaluated. However, we decided to consider only BAL samples derived from patients who started treatment within 7 days prior to BAL sampling in order to assess the potential influence of treatment on cytokine levels. Due to the limited sample size, we restricted the analysis to tocilizumab, steroids and antiviral treatments (remdesivir, lopinavir/ritonavir) also including in this last group, hydroxychloroquine. Figure 4 shows that we observed a significant difference between different groups only in the case of IL6: patients treated with steroids had lower BAL IL6 compared to those who received tocilizumab (p<0.1) or antiviral drugs (p<0.05) (Fig. 4a).

**Figure 4.**
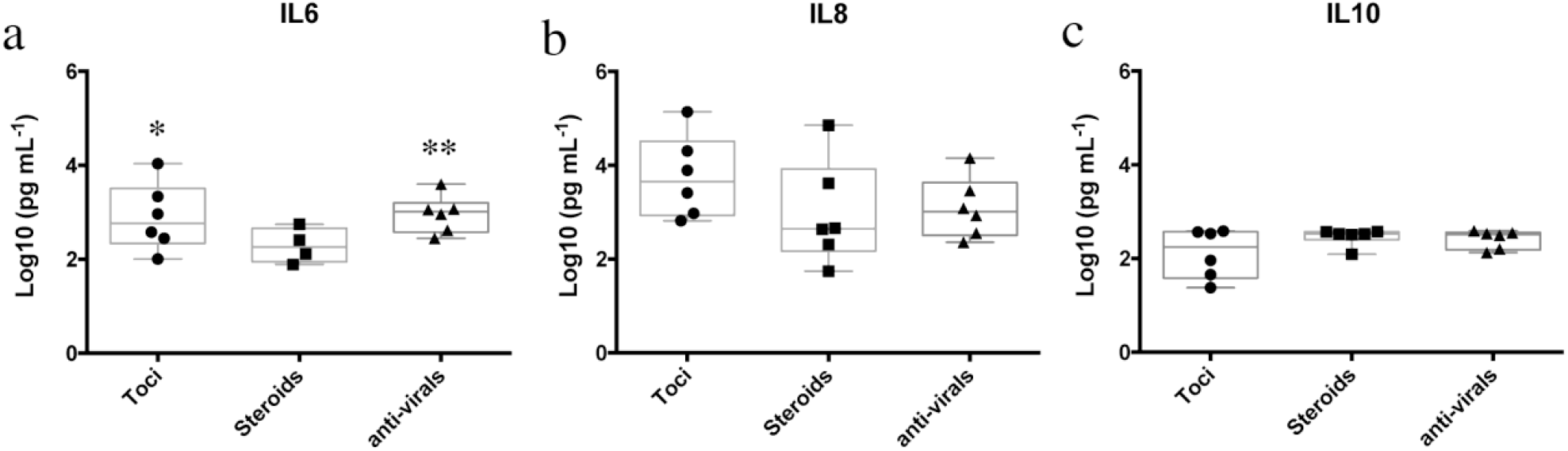
Differences between cytokines after tocilizumab, steroids, and anti-viral drugs treatment. Hydroxychloroquine has been added in the anti-viral group. Data are represented as median (IQR). *p < 0.1 *vs*. steroids; **p < 0.05 *vs*. steroids.

Finally, we investigated whether BAL cytokines correlated with differential cell counts quantified previously (Table 3) or between each other (Table 4), considering all collected BAL samples. We found that IL6 and IL8 correlated directly with neutrophils (Table 3 and Fig. S4b) and inversely with macrophages percentage (Table 3 and Fig. S4a). We observed that the level of IL8 also correlated inversely with lymphocytes percentage (Table 3). Analyzing the correlation between cytokines, levels of IL6 and IL8 were correlated between each other, while IL10 correlated inversely with IL8 (Table 4). These correlations between cytokines are clear in two representative cases of patients subjected to repeated bronchoscopies (Fig. S5).

**Table 3.**
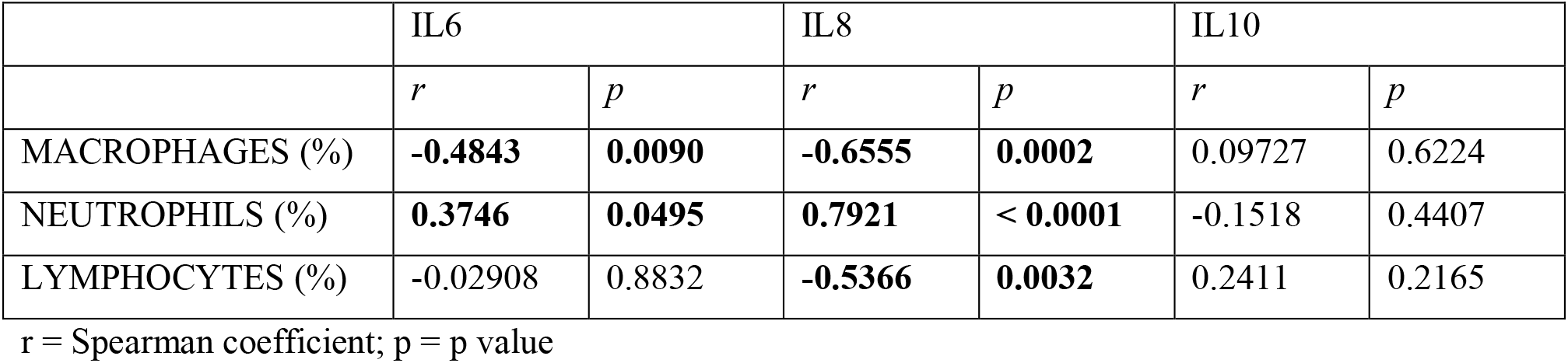
Correlation analysis of entire BAL collected between cell populations and cytokines.

**Table 4.**
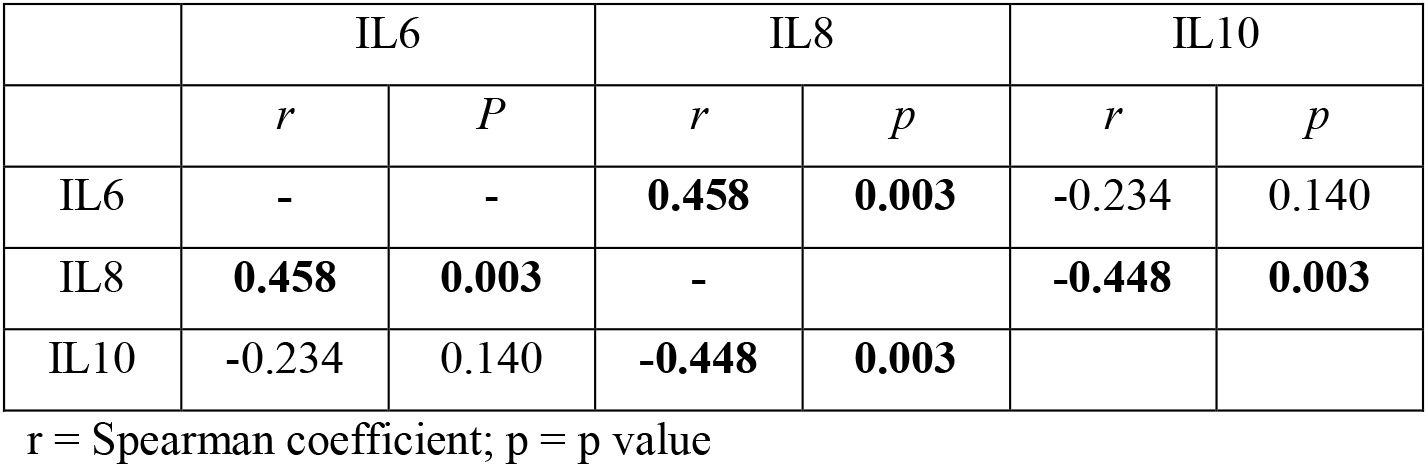
Correlation analysis between cytokines quantified in BALs overall.

|  | IL6 |  | IL8 |  | IL10 |  |
| --- | --- | --- | --- | --- | --- | --- |
|  | *r* | *P* | *r* | *p* | *r* | *p* |
| IL6 | - | - | **0.458** | **0.003** | -0.234 | 0.140 |
| IL8 | **0.458** | **0.003** | - |  | **-0.448** | **0.003** |
| IL10 | -0.234 | 0.140 | **-0.448** | **0.003** |  |  |

r = Spearman coefficient; p = p value

For patients from whom we collected IL6 plasma and BAL quantification sampling on the same day (20 from ICU and 4 from IMW) we analyzed the correlation and observed a direct correlation (Spearman coefficient = 0.53 with p<0.01) (Fig. S6).

## Discussion

Up to now, there has been little information about the deep lung inflammatory status in SARS-CoV-2 patients. In the present study we analyzed the broncho-alveolar inflammatory environment of patients admitted to the ICU and IMW of two hospitals situated in the epicenter of the Italian epidemic outbreak. In particular, we evaluated cell differentials, cell activation by morphological features, and cytokines assessment on retrieved BAL fluids. Among cytokines, we decided to focus our attention on IL6, a well-known inflammatory cytokine that has been used as a marker of macrophages activation in the peripheral blood of COVID-19 affected patients [13]; IL8, a pro-inflammatory CXC chemokine that exerts specific chemotactic and activating functions on neutrophils; and IL10, an anti-inflammatory and immunoregulatory cytokine, produced by several immune effectors, whose role in SARS-CoV-2 infected patients is still in debate [9, 14].

In severe ICU COVID-19 patients, the alveolitis was associated with hyperactivation of innate effectors, macrophages and neutrophils, that showed pseudopodia and cell-to-cell contacts (Figure 2a and b). Moreover, a large number of cytopathic cells with vacuolization, osmiophilic bodies, loss of integrity of both nuclear and plasma membranes and cell fragmentation were present (Fig.1, 2 and S2). Immunostaining with anti-viral capsid and spike antibodies specifically immunoreacted with BAL cells, mostly cytopathic ones (Fig. 1e,f and Fig. 2c and d).

High percentage of neutrophils in ICU patients BAL in contrast to IMW patients were founded (Fig. S1a and b). The high infiltration of neutrophils inside the alveolar space has already been assessed in severe COVID- 19 patients, but only after autopsy study [15]. Herein, we quantify the neutrophil percentage in the alveolar space from mild to severe stages of COVID-19, confirming the literature.

Regarding lymphocytes, we observed a strongly and significantly reduced percentage in ICU patients as compared to those in the IMW (Fig. S1), corroborating findings recently reported by Liao and colleagues who analyzed BAL samples from 3 severe and 3 mild SARS-CoV-2 patients. The authors showed a decreased lymphocyte counts in the three severe patients with respect to the mild ones [10]. Thus, we have added another insight regarding the impairment of adaptive immune response to COVID-19 reported in peripheral blood, suggesting the same behavior also at the lung level [6, 8, 16, 17].

Concerning the three analyzed cytokines, we demonstrated that patients admitted to the ICU are characterized by significant high levels of the two pro-inflammatory cytokines, IL6 and IL8, with respect to IMW patients (Fig. 3a,b). This result is sustained by previous reports obtained for Middle East Respiratory Syndrome (MERS) pneumonia patients, demonstrating a severe down-regulation of Th2, and inadequate Th1 immune response in respiratory specimens together with high expression levels of inflammatory cytokines IL-1α and IL-1β and IL-8 (CXCL8) [18]. Furthermore, a recent study about single-cell RNA profiles of alveolar macrophages from 3 SARS-CoV-2 patients identified a prevalence of macrophages expressing FCN1 and several pro-inflammatory genes (CCL2, CCL3, CCL5, IL-8, CXCL9, CXCL10 e CXCL11), while anti-inflammatory macrophages, and in particular macrophages involved in surfactant turnover, were significantly reduced in severe COVID-19 patients [10].

The study herein also demonstrated that upon analyzing survivors in comparison to non-survivors, we found significantly lower IL6 (Fig. 3c) and IL8 (Fig. 3d) levels in the survivor group with respect to non-survivors. The analysis of treatment effect on BAL inflammation, limited to a small number of ICU patients, showed that in patients submitted to a steroid course IL6 was lower than in those treated with tocilizumab or anti- viral drugs (Fig. 4a). It should be noted, however, that tocilizumab, by acting at a receptor level, does not interfere with IL6 release, but rather exerts its action on the IL6-driven inflammatory cascade. The same trend towards a decrease in IL6 was also observed for IL8, which did not reach statistical significance (Fig. 4b). Thus, taking together all our findings, we can suggest that the burden of pro-inflammatory cytokines IL6 and IL8 in deep lung is not only related to the severity of the disease but might also be associated to the outcome. Steroids seem to be able to limit the release of IL6 and IL8 while not significantly affecting IL10 levels (Fig. 4c). This last result is supported by a clinical trial called “RECOVERY”, which reported that dexamethasone reduced deaths by one-third in ventilated patients [19].

The role of IL10 in COVID-19 infection is a matter of ongoing debate. Increased serum levels of IL10 have been described in severe SARS-CoV-2 patients with respect to those affected by mild disease. Moreover, authors have demonstrated a direct correlation with IL6 serum levels, suggesting that both cytokines could be used as predictors of higher risk of disease deterioration [20]. Thus, despite its well-known anti-inflammatory role, an exaggerated release of IL10, together with other pro-inflammatory factors from activated macrophages is integrated in the context of the cytokine storm and considered a marker of disease severity [21]. In the present study, however, there was no significant variation in IL10 BAL levels either in relation to disease severity or according to survival (Fig. 3). In addition, no significant correlation was found between IL10 and IL6 in BAL, while only a negative and significant correlation with IL8 levels was present (Table 4). Thus, on the basis of the present study, a role of BAL IL10 as a specific disease marker cannot be inferred.

We are aware that this study has some limitations. We performed these preliminary analyses on limited sample size given the high risk of infection of the health care personnel. This limited our possibility to make further analyses, such as cell surface activation markers assessment by flow cytometry on macrophage or lymphocytes. Another limitation is the lack of paired assessment of cytokines in the peripheral blood due to the absence of stored serum samples from acute COVID19 patients in our institutions. Finally, we know that the infectious complications registered (Table 1) might have influenced cell differentials and cytokine levels however, the percentage of bacterial and fungal co-infection was very high in all patients in ICU without a significant difference between survivors and non-survivors.

As a result of our study we wish to highlight the possible crucial role of IL8 in COVID-19 infection at the lung level. This cytokine showed higher values among ICU compared to IMW patients (Fig. 3a and b) and was associated to a negative outcome (Fig. 3d). It is known that IL8 acts as a chemoattractant and activating factor of neutrophils, in fact we assessed a direct correlation between IL8 and neutrophils percentage in BAL (Table 3 and Fig. S4b). Hence, we can infer that the release of IL8 represents a crucial step of SARS-CoV-2 pathogenesis and its pathway might represent a possible target of future intervention.

## Data Availability

Data will be available in anonymized form by request to the corresponding author for research purposes, after approval by the institutional ethical committee, and after approval by all co-authors

## Acknowledgements

Monica Concardi: Center for Inherited Cardiovascular Diseases, IRCCS Policlinico San Matteo Foundation, University of Pavia, Pavia, Italy

Marco Manstretta: Research Laboratory of Lung Diseases, IRCCS Policlinico San Matteo Foundation, University of Pavia, Pavia, Italy.

Carola Marioli: Pneumology Unit, IRCCS Policlinico S. Matteo Foundation, Pavia, Italy

